# Uncovering distinct motor development trajectories in infants during the first half year of life

**DOI:** 10.1101/2025.02.06.25321826

**Authors:** Riley C Elmer, Moon Sun Kang, Beth A Smith, Ran Xiao

## Abstract

Infants undergo significant developmental changes in the first few months of life. While some risk factors increase the risk of developmental disability, such as preterm birth, the developmental trajectories of infants born pre-term (PT) and full-term (FT) present with individual variability. This study aims to investigate whether the utilization of data-driven unsupervised machine learning can identify patterns within groups of infants and categorize infants into specific developmental trajectories. Thirty-four infants, 19 FT and 15 PT, were assessed with the gross and fine motor subscales of the Bayley Scales of Infant and Toddler Development, version III (BSID-III) monthly for 2-5 visits between the ages of 1 and 6 months. Latent class growth analysis (LCGA) models were adopted to identify clusters of motor developmental trajectories during this critical time. Based on statistical significance, the linear, 2-class trend was selected as the best-fitting model for both gross and fine motor trajectories. Within this, LCGA reveals 2 developmental trends with varying beginning scores and developmental rates, including the low-baseline slow-growth (LBSG) subgroup, and the high-baseline fast-growth (HBFG) subgroup, with age (adjusted for prematurity) being equally distributed across both subgroups. Both subgroups, HBFG and LBSG, had a combination of infants born FT and PT (55% FT in HBFG, 56% FT in LBSG), supporting that preterm birth alone may not sufficiently categorize an infant’s developmental trajectory. The later BSID-III gross motor score showed marginal difference between groups (p = 0.062). Similarly, the fine motor model displayed a mixture of both infants born FT and PT (68% FT in HBFG, 40% FT in LBSG). In this case, the late motor composite BSID score was different between groups (p = 0.04). Our study uses a novel approach of LCGA to elucidate heterogeneous trajectories of motor development for gross and fine motor skills during the first half of life and offers potential for early identification of subgroup membership. Furthermore, these findings underscore the necessity for individualized risk assessments and intervention strategies tailored to individual needs. Ultimately, further validation of these models may provide usefulness in uncovering distinct motor development trajectories in infants.

**Author Note:** The authors would like to thank the infants and families who participated in these study, and Katy Kelley, Carolina Panceri, Judy Zhou, and Dana Fine for assisting with recruitment and data collection. Thank you to Eisner Health, Children’s Hospital Los Angeles, and Ventura County Medical Center for assistance with recruitment.

The authors declare that the research was conducted in the absence of any commercial or financial relationships that could be construed as a potential conflict of interest. The studies secondarily analyzed in this work were supported by grants from the Bill & Melinda Gates Foundation [OPP1119189] (PI: B.A.S.) and the Eunice Kennedy Shriver National Institute of Child Health & Human Development of the National Institutes of Health under Award Number R03HD096137 (PI: Smith). The content is solely the responsibility of the authors and does not necessarily represent the official views of the National Institutes of Health. Study data were collected and managed using REDCap electronic data capture tools hosted at the Southern California Clinical and Translational Science Institute at the University of Southern California NIH grant UL1TR001855 (PI: T. Buchanan). REDCap (Research Electronic Data Capture) is a secure, web-based application designed to support data capture for research studies, providing (i) an intuitive interface for validated data entry; (ii) audit trails for tracking data manipulation and export procedures; (iii) automated export procedures for seamless data downloads to common statistical packages; and (iv) procedures for importing data from external sources.

## Uncovering distinct motor development trajectories in infants during the first half year of life

Humans undergo significant developmental transitions from two to four months of life (Hadders-Algra, 2022). During this time, infants’ sensory systems develop to promote increased interaction with their environment. These experiences contribute to necessary motor skill development. Motor skills during infancy are measurable milestones and are positively correlated with later cognitive and language skill performance (Bolger et al., 2021; Collett et al., 2019; Ghassabian et al., 2016; Gonzalez et al., 2019; Holloway & Long, 2019; Sansavini et al., 2021). For example, fine motor skill performance can serve as a predictor of other developmental outcomes, such as school readiness (Grissmer et al., 2010; Iverson, 2010). Tracking motor skill acquisition can provide unique insights into an infant’s developmental trajectory. Examining a cohort of infants may elucidate patterns or characterizations that can inform future targeted research questions and hypotheses.

Early milestones of motor development that emerge in the first few months of life, such as reaching, are especially valuable in research and clinical settings. It remains difficult to extract predictive information for individuals, however, as significant variability exists between infants. Predicting developmental delay or disorders can be especially challenging. These challenges are among the reasons that many disorders are not diagnosed until late infancy or early childhood (Brown et al., 2020), despite the increase in the prevalence of developmental disability since the late 2000s (Zablotsky et al., 2019).

Extracting patterns of developmental trajectory provides key insights for both preterm (PT) and full-term (FT) infant populations. As gestational age at birth decreases, the risk of developmental disability increases exponentially (Kerstjens, de Winter, et al., 2012). In PT infants, the prevalence of motor delay or disability is significantly higher than those born full-term (Bos et al., 2013; Ko & Lim, 2023). Early identification of one’s motor development trajectory patterns may support earlier, targeted intervention. Some evidence suggests that motor-based therapeutic intervention as early as 3 months of age may critically influence outcomes (Hughes et al., 2016). Unfortunately, standards on if and when to begin treatment or developmental support vary widely (Hee Chung et al., 2020). These discussions derive largely from the heterogeneity of human development. Rather than relying on individual risk factors for developmental delay or disorder, a multifactorial system is most effective for elucidating developmental trajectory. An alternative approach to addressing these challenges is to track the motor development of a group of infants varying in gestational age at birth, then perform unsupervised statistical approaches to subdivide the sample without assumptions about risk factors. This method allows for the complexity and heterogeneity of a population while uncovering potential unforeseen influences. Identifying patterns of early development is a critical predecessor to targeted clinical care. Characterizing groupwise longitudinal development allows us to explore data-driven patterns or trajectories that may emerge and inform future predictive models.

Developmental assessments are used in clinical and research settings to assess a child’s motor, language, and cognitive skill. One of these measures is the Bayley Scales of Infant and Toddler Development (BSID), an assessment that tracks progression relative to their age-matched peers (Bayley, 2006). During the BSID, infants are presented with tasks that relate to memory, language, communication, fine and gross motor skills, and more. Each of these subcategories are scored individually and report the number of related tasks the infant is able to complete. These values are then normalized to report percentile information that can inform clinical care. Within the BSID-III, motor development is parsed into two categories: gross motor and fine motor. Gross motor skills are classified as movements that involve large muscle groups, and fine motor skills are generally associated with smaller muscle groups.

The BSID describes tendencies of developmental achievement at a point in time but does not predict an infant’s developmental trajectory. This time-stamped measure contributes to a limited conceptualization of how infants will change over time. However, statistical methods can incorporate BSID information to provide insights into the longitudinal change of an infant’s development. The Latent Class Growth Analysis (LCGA), a derivative of Growth Mixture Modeling (GMM), serves as a potent tool for delineating distinct groups within longitudinal data based on their developmental trajectories (Lanza, 2016). LCGA has been widely adopted in developmental research, with various published studies establishing its efficacy. For instance, Heineman et al. utilized LCGA on the Infant Motor Profile (IMP) at 4, 10, and 18 months to explore the links between early motor development and cognitive functions at age four, elucidating that even amidst individual variation, a strong relationship exists between motor development and cognition (Heineman et al., 2018). Similarly, Valla et al. conducted LCGA on data from the Ages and Stages Questionnaire (ASQ) collected at multiple time points—4, 6, 9, 12, 16, and 24 months—identifying three distinct developmental profiles in gross and fine motor skills: high stable, U-shaped, and late bloomers (Valla et al., 2017). These findings highlight LCGA’s capacity to reveal insightful patterns in developmental research, albeit with relatively broad age intervals. Notably, there is a gap in understanding the trajectory of how the earliest intentional fine and gross motor milestones, such as reaching, emerge.

This study aims to address this gap by analyzing monthly motor developmental trajectories (BSID-III gross and fine motor skill subscales) of infants born preterm and full-term through LCGA. By characterizing distinct developmental patterns, this research will enhance our understanding of the variability in early motor development, inform future research questions regarding motor trajectories across the first half year of life, and provide foundational information into the mechanisms of individualized developmental support.

## Method

### Study Design

This study performed secondary analyses on data from two previous separate, published studies. This section outlines the extraction criteria for this study and the study design in the original publications. Both studies were conducted within the same research lab, providing samples to elucidate potential trajectories that may emerge from FT and PT infants.

#### Study 1

The data for the infants born full term and for part of the preterm infants were extracted from a study previously published by this lab (Hooyman et al., 2018). This study was approved by the Institutional Review Board of the University of Southern California, and informed consent was obtained from a parent or legal guardian before participation. The study was conducted at the University of Southern California Health Science Campus. The inclusion criteria for infants born full term were singleton births at more than 38 weeks gestational age, as well as being free of any visual, orthopedic, or neurological impairment. Additionally, infants were excluded if they scored at or below the 5^th^ percentile on the BSID-III. Infants born preterm were included based on criteria set for high-risk infant follow-up services in the State of California or were born before 36 weeks of gestation (Services, 2020). The target sample of the study was infants at increased risk for developmental disabilities, but every infant except one was born preterm. Infants with unstable medical conditions were excluded (Bayley, 2006). Infants were enrolled in the study before they demonstrated skilled reaching movements and were assessed across the time period that reaching emerged.

#### Study 2

Additional data from infants born preterm were extracted from a separate, published study from this lab (Nishiyori et al., 2021). This study was approved by the Institutional Review Board of the University of Southern California, and informed consent was obtained from a parent or legal guardian before participation. The study was conducted at the University of Southern California Health Science Campus. In this study, infants were eligible to enroll if their gestational age at birth was less than 32 weeks. Infants with unstable medical conditions were excluded. Infants were enrolled in the study before they demonstrated skilled reaching movements and were assessed across the time period that reaching emerged.

### Data Collection Procedures

#### Bayley Scales of Infant and Toddler Development III (BSID-III)

The BSID-III is a standardized assessment that can be utilized from 16 days to 42 months of age and takes between 30-70 minutes to administer (Bayley, 2006). During the BSID-III, infants were presented with tasks that relate to memory, language, communication, fine and gross motor skills, and more.

Regarding gross motor skills, the BSID, version 3 (BSID-III) Administration Manual states, “Items measure static positioning (e.g., sitting, standing); dynamic movement, including locomotion and coordination; balance; and motor planning” (Bayley, 2006). For fine motor skills, the BSID-III describes that “these items measure” skills related to visual tracking, reaching, object manipulation, and grasping” (Aylward, 2019; Bayley, 2006; Brown et al., 2020; Burakevych et al., 2017; Kerstjens, de Winter, et al., 2012; Zablotsky et al., 2019).

The infants received a raw score based on task performance. The raw scores were then used to create a scaled score through inferential norming. Statistical measures such as the mean, standard deviation (SD), and skewness were calculated for each age group. They were then used to fit a nonlinear model based on theoretical expectations and growth curve patterns (Aylward, 2019). These norm-referenced values were used to create Gaussian distributions for each age group, thus creating percentiles for each category. The standard scores are a sum of the relevant scaled scores for each category and adjusted so the mean value is 100, with a standard deviation of 15 (Burakevych et al., 2017). The BSID scores reported in this study are in both the raw scores (Fine Motor and Gross Motor) and standard scores (Motor Composite) format.

For Study 1, the cognitive, motor, and language assessments of the BSID-III (Bayley, 2006), were collected monthly between infant ages 1 to 6 months, with 2-5 visits for each participant. Infants were primarily assessed in their own homes, with few being assessed in the lab space. The BSID-III was conducted by a trained research associate. The administrator was not blind to the infant’s gestational age at birth. Anthropometric and demographic data were also collected at each visit. Additionally, electroencephalography recordings, synchronous video data, and wearable motion sensor data were collected during a reaching skill assessment but were not analyzed in this study.

For Study 2, anthropometric, demographic, and BSID-III scores were collected monthly from infants between ages 1-6 months corrected age, with visits ranging from 1-5 per infant. The BSID-III was conducted by a trained research associate. Assessments were completed either in the participant’s home, or at the research director’s lab at the University of Southern California. Electroencephalography (EEG) data, synchronous video data, and wearable motion sensor data were collected from these infants but were not analyzed in this study and will not be discussed further.

### Participants

There was a combined total of 46 infants who participated in both studies, including 22 FT and 24 PT. FT is defined as infants born with gestational age > 37 weeks. PT is defined as being born before 37 weeks of gestational age. We excluded 1 PT and 2 FT infant data from further analysis as their starting age in the study exceeded 6 months of age. We further excluded 8 PT and 1 FT infants when they participated in less than 3 sessions resulting in too few temporal data points for analyzing the developmental trends. One or two data points are insufficient in informing the shape and potential acceleration/deceleration patterns within each trajectory. The detailed inclusion/exclusion criteria are listed in **Fig. 1**. The final analysis of this study included 34 infants, with 19 FT and 15 PT. An average of 3.76 (SD: 0.82) visits were collected from each of the remaining participants. For both Study 1 and Study 2, visits were spaced out monthly. However, infants did not begin and end the study at the same monthly timepoints. The largest amount of data was collected within the 3–5-month age range. The distribution of the infant’s age throughout the data collection can be found in **Fig. 2**. The gross and fine motor trends of both FT and PT populations across visits are shown in **Fig. 3**. There were 22/34 (∼65%) females in the overall sample, with 11/19 (∼58%) female FT infants and 11/15 (∼73%) female PT infants. For additional information on demographic characteristics, see **Table 1**.

**Figure 1.**
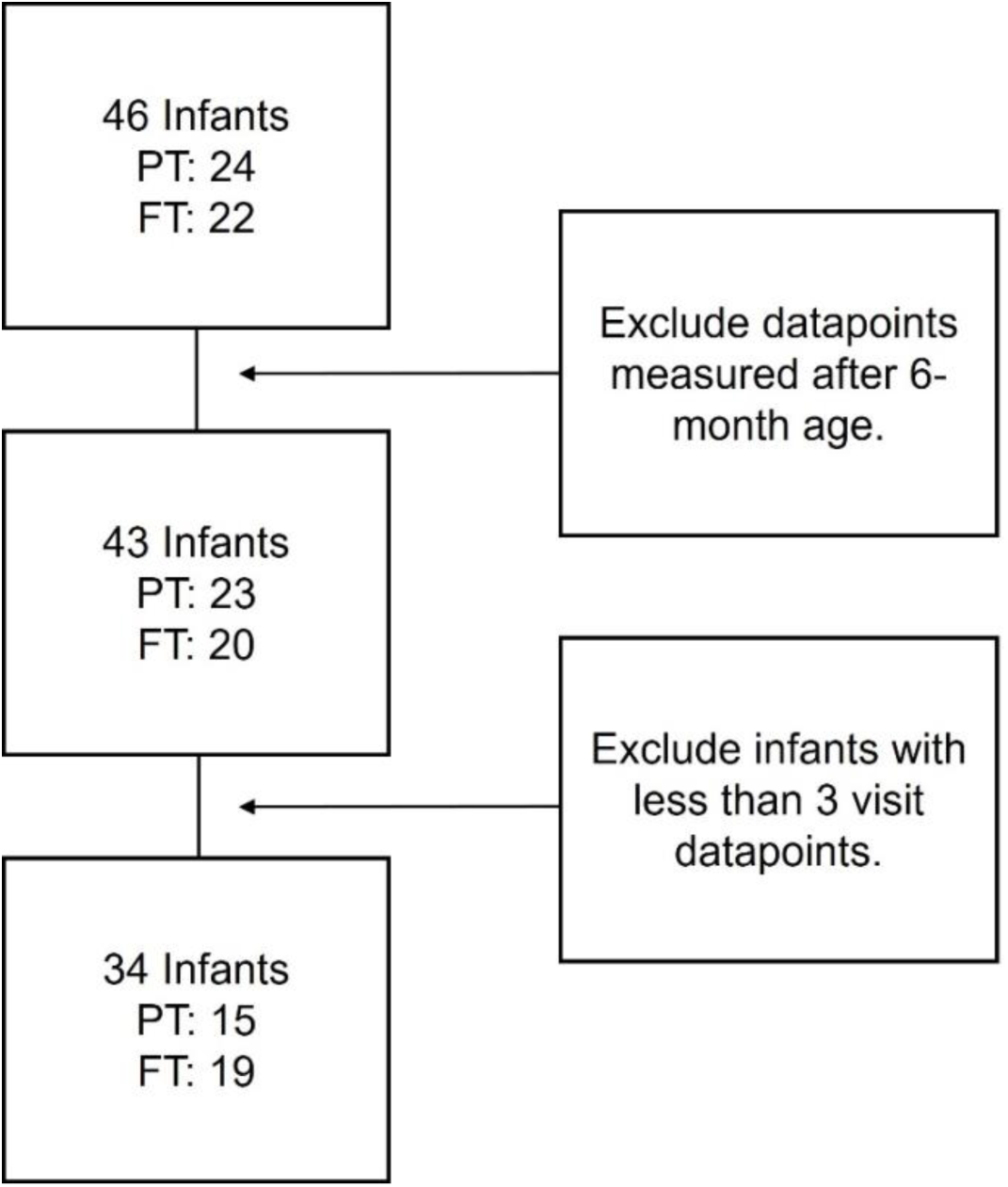
Flow diagram demonstrating the participant inclusion criteria and final participant number. PT = Infant born preterm, FT = infant born full term.

**Figure 2.**
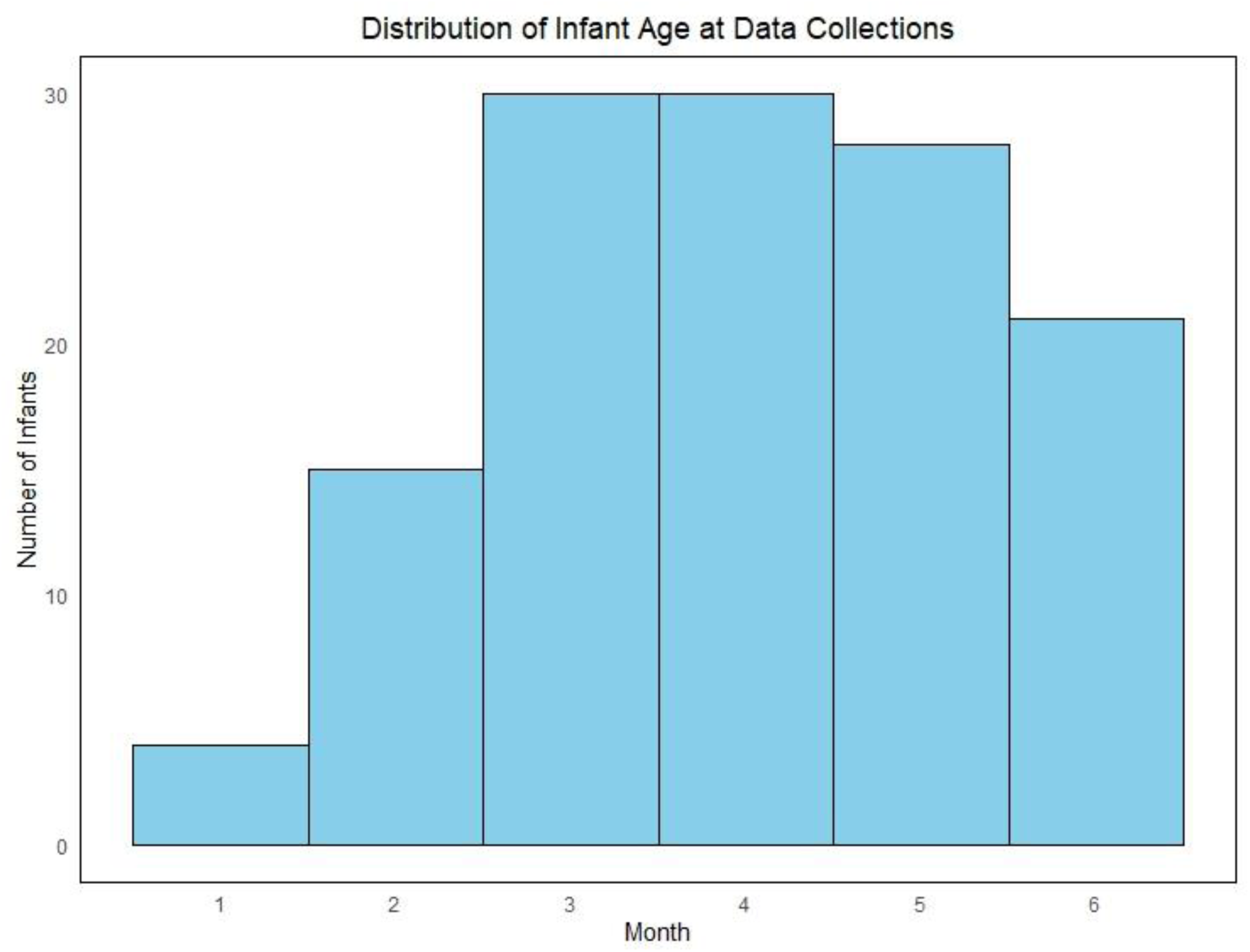
Histogram displaying the number of infants for which developmental data was collected at each of the first six months of life.

**Figure 3.**
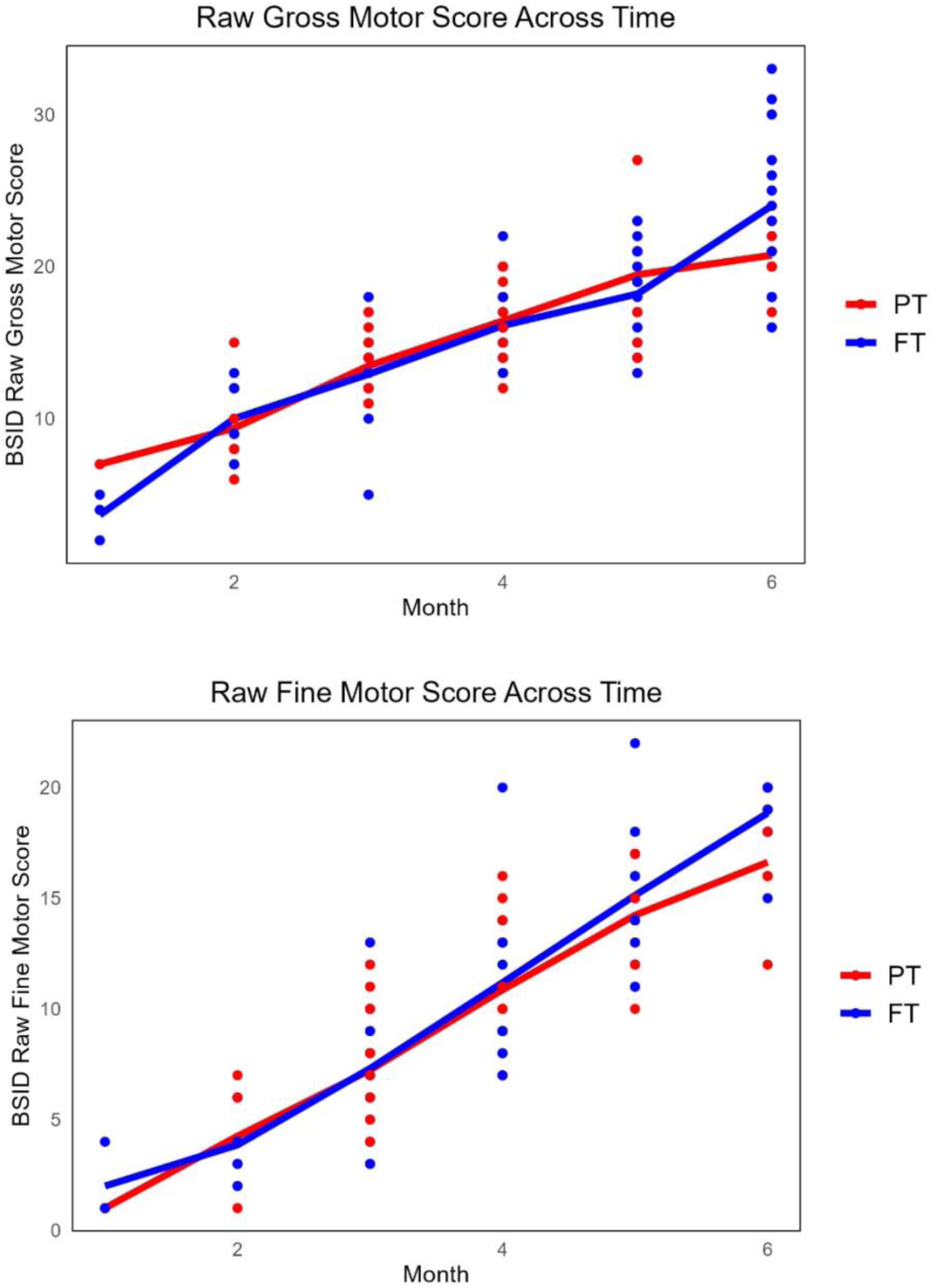
Scatterplot showing raw BSID Gross and Fine Motor Scores. Red = Participants born preterm (PT), Blue = Infants born full term (FT), BSID = Bayley Scales of Infant and Toddler Development, version III. X-axis is age in months, adjusted for preterm birth.

**Table 1.**
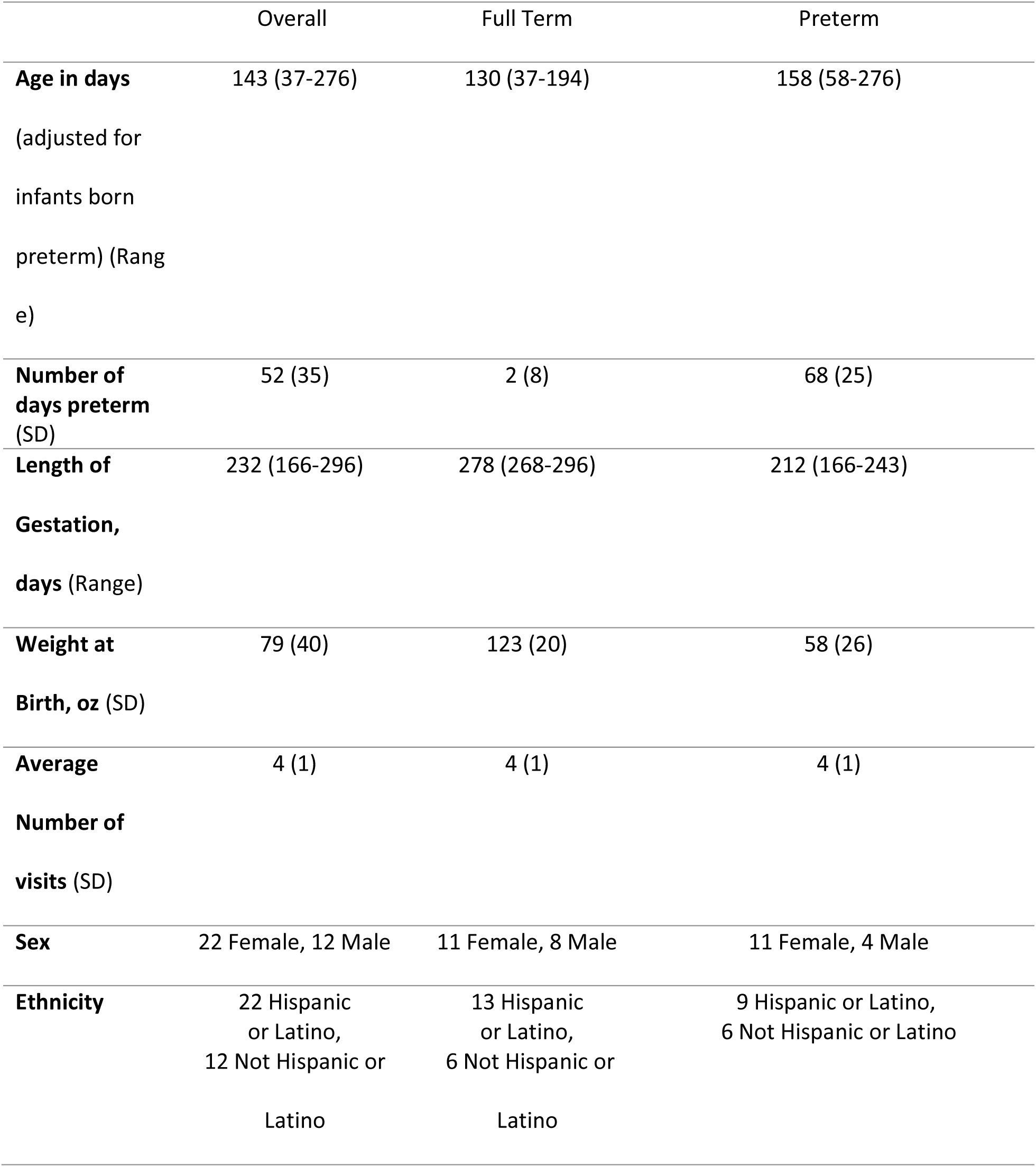
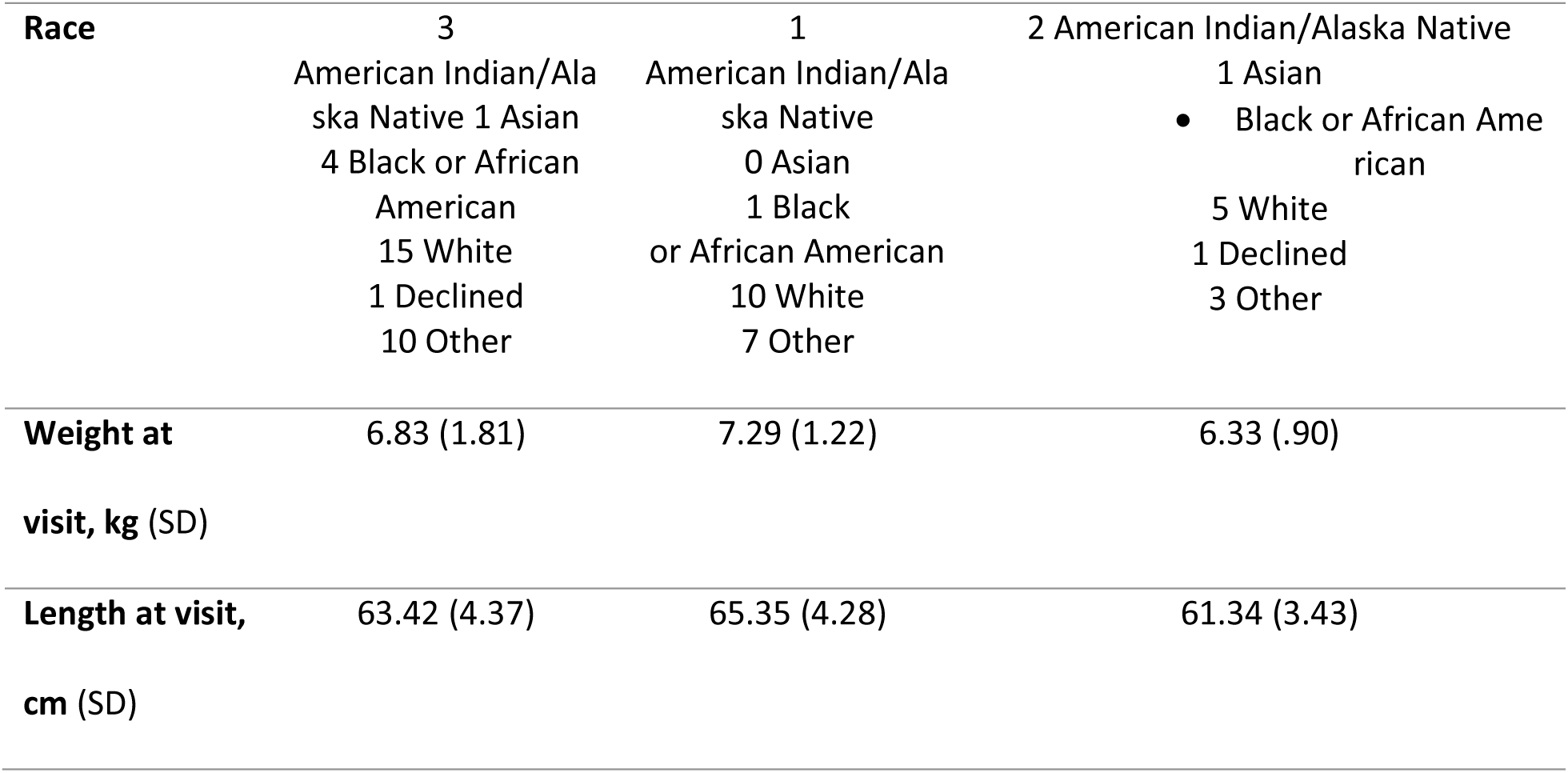
Demographic information of infant participants.

### Data Analysis

#### Identifying Developmental Trajectories Through Latent Class Growth Analysis (LCGA)

In this study, we utilized LCGA, a data-driven approach that identifies latent classes, or subgroups with distinct patterns, within the longitudinal data. LCGA was chosen because of its capability to handle missing data through full information maximum likelihood estimation, which utilizes all available data and assumes data are missing at random. This effectively mitigates the issue of longitudinal studies of this nature, where infants may have variable starting points, missing assessments in certain months, or early drop-off. We explored models with 2, 3, 4, and 5 classes to determine the most appropriate number of subgroups, based on a combination of statistical measures described in detail below.

To accommodate different developmental trends in fine and gross motor skills, we assessed the data using both linear and non-linear models. Linear trends were delineated using the ‘hlme’ function in R, which implements hierarchical linear mixed effects modeling. For non-linear trends, we employed the ‘lcmm’ package with splines as the linkage function, allowing for flexible curve fitting.

Optimal model selection was achieved through a systematic grid search technique, which was rigorously repeated 100 times to enhance the likelihood of identifying the global optimum rather than merely a local one. This approach aims to ensure more stable and reliable results by extensively exploring the parameter space. Each model fitting was allowed a maximum of 100 iterations to achieve optimal convergence.

Statistical selection of the best model was guided by the Akaike Information Criterion (AIC) and the Bayesian Information Criterion (BIC). AIC seeks to identify the model that best explains the data with the fewest parameters, with a lower score indicating a better balance of fit and complexity. Conversely, BIC imposes a stricter penalty on model complexity, making it particularly valuable for larger datasets.

By evaluating both AIC and BIC scores across models with varying assumptions and numbers of classes, and with consideration of sample distributions across different subgroups, we determined the best-fitting models for both fine and gross motor skill scales.

#### Comparison Between Subgroups

Given that LCGA focuses on identifying subgroup-level trajectories rather than individual trends at the level of each infant, we characterized differences among the latent subgroups by segmenting the data by monthly age. This allows for the extraction of potential patterns or characteristics that could inform the subgroup trajectories. Specifically, median motor scores from Months 1 to 3 were categorized as early scores, while those from Months 4 to 6 were classified as late scores. This approach mitigates the limitation of varying starting ages across participants and improves sample coverage from both subgroups for statistical comparisons. A two-sample Wilcoxon Rank Sum Test was then conducted to compare early scores between the two subgroups, followed by a comparison of late scores. The same process was applied to both fine and gross motor raw scores. Given the relatively small sample size in this study, we align with the approach discussed in (Olsson-Collentine et al., 2019; Pritschet et al., 2016), which highlights the ongoing shift toward reporting not only statistical significance at p < 0.05 but also marginal significance within the range of 0.05 < p < 0.10. This approach aims to identify meaningful patterns of significance in trajectories while reducing the risk of overlooking potentially important differences in a relatively small sample.

## Results

### Phenotyping Developmental Trajectories of Gross Motor Scales

Table 2 shows the LCGA analysis results for gross motor scales. For the gross motor models, the linear, 2-class model was identified as the best fit, supported by statistical measures (AIC = 627.34, BIC = 636.49) and considerations of sample distribution. Although some models with higher class numbers showed slightly lower AIC and BIC values, they often resulted in a single participant being assigned to a unique subgroup. In the selected 2-class linear model for gross motor skills, 52.94% of participants were classified into Class 1 and 47.06% into Class 2. In contrast, the 3-class, 4-class, and 5-class models each categorized only one infant into an individual class, further highlighting the risk of overfitting. In addition, the linear model was chosen over the nonlinear one, due to a balance of statistical measures, model complexity (the number of parameters (NPM) in the 2-class linear model is 6, whereas it is 11 in the nonlinear model), and sample distribution. Although the small sample size may be considered a limitation, the chosen model’s lower AIC and BIC values, along with better sample distribution, reinforce the appropriateness of a 2-class model. This will be further discussed in the Discussion section.

**Table 2.**
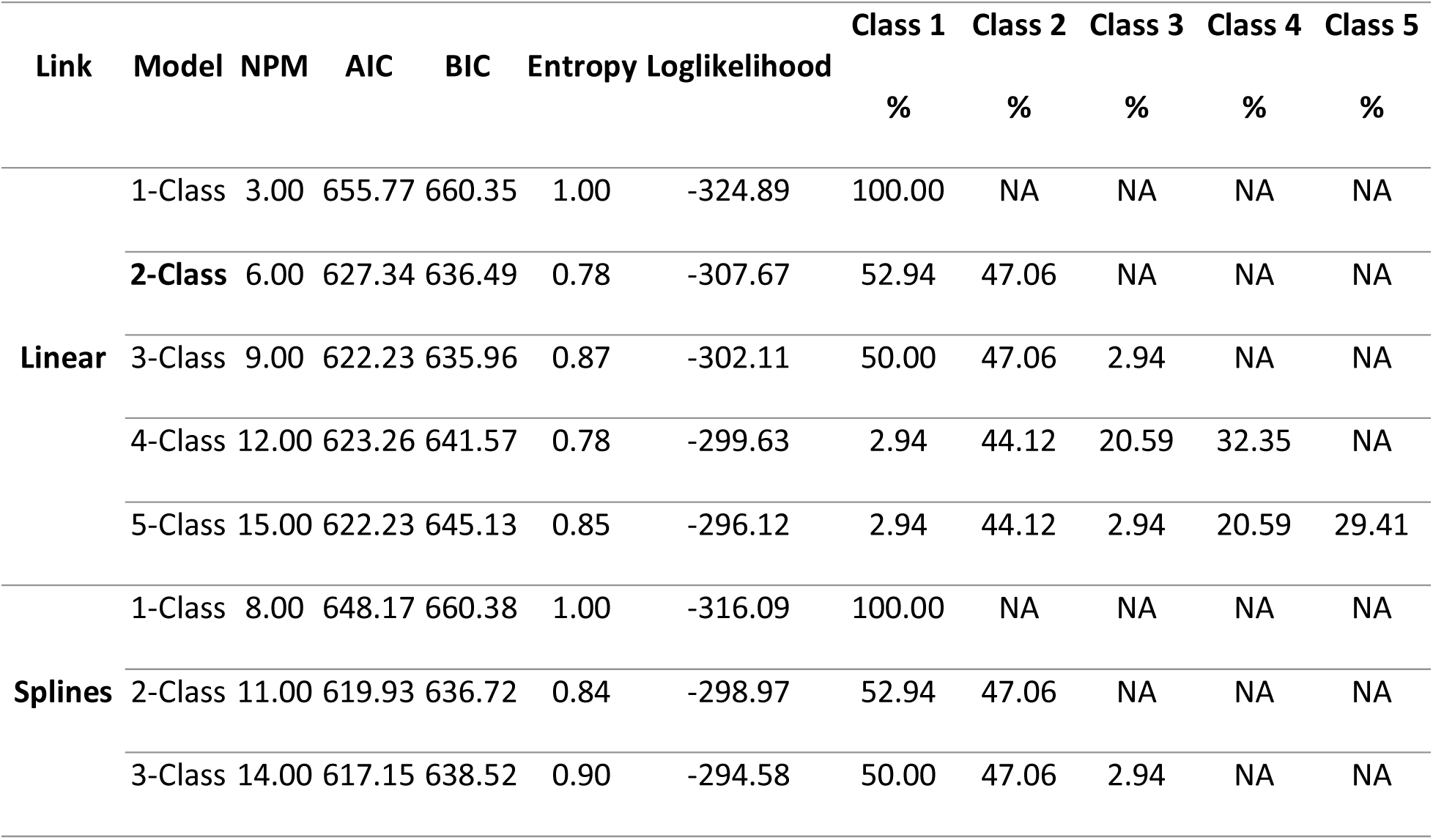
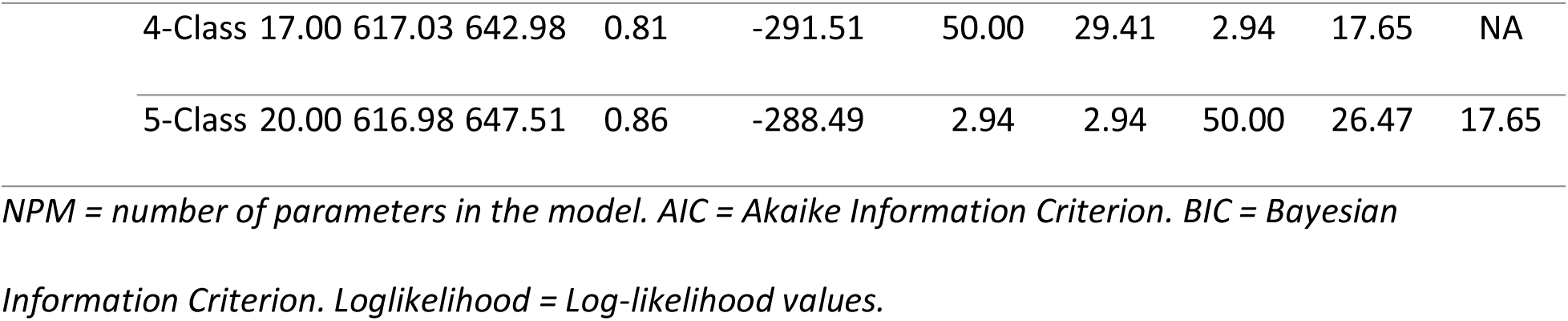
Latent Class Growth Analysis results for gross motor scales. NPM = number of parameters in the model. AIC = Akaike Information Criterion. BIC = Bayesian Information Criterion. Loglikelihood = Log-likelihood values.

| Link | Model | NPM | AIC | BIC | Entropy | Loglikelihood | Class 1 | Class 2 | Class 3 | Class 4 | Class 5 |
| --- | --- | --- | --- | --- | --- | --- | --- | --- | --- | --- | --- |
|  |  |  |  |  |  |  | % | % | % | % | % |
| Linear | 1-Class | 3.00 | 655.77 | 660.35 | 1.00 | -324.89 | 100.00 | NA | NA | NA | NA |
|  | 2-Class | 6.00 | 627.34 | 636.49 | 0.78 | -307.67 | 52.94 | 47.06 | NA | NA | NA |
|  | 3-Class | 9.00 | 622.23 | 635.96 | 0.87 | -302.11 | 50.00 | 47.06 | 2.94 | NA | NA |
|  | 4-Class | 12.00 | 623.26 | 641.57 | 0.78 | -299.63 | 2.94 | 44.12 | 20.59 | 32.35 | NA |
|  | 5-Class | 15.00 | 622.23 | 645.13 | 0.85 | -296.12 | 2.94 | 44.12 | 2.94 | 20.59 | 29.41 |
| Splines | 1-Class | 8.00 | 648.17 | 660.38 | 1.00 | -316.09 | 100.00 | NA | NA | NA | NA |
|  | 2-Class | 11.00 | 619.93 | 636.72 | 0.84 | -298.97 | 52.94 | 47.06 | NA | NA | NA |
|  | 3-Class | 14.00 | 617.15 | 638.52 | 0.90 | -294.58 | 50.00 | 47.06 | 2.94 | NA | NA |

|  |  |  |  |  |  |  |  |  |  |  |
| --- | --- | --- | --- | --- | --- | --- | --- | --- | --- | --- |
| 4-Class | 17.00 | 617.03 | 642.98 | 0.81 | -291.51 | 50.00 | 29.41 | 2.94 | 17.65 | NA |
| 5-Class | 20.00 | 616.98 | 647.51 | 0.86 | -288.49 | 2.94 | 2.94 | 50.00 | 26.47 | 17.65 |

The developmental trajectories of each class can be found in **Fig. 4**. The two classes elucidate two trajectories hereto differentiated as the high-baseline fast-growth (HBFG) subgroup, Class 1, and the low-baseline slow-growth (LBSG) subgroup, Class 2. The HBFG subgroup is characterized by high initial raw gross motor scores and a more rapid developmental rate across visits, while the LBSG subgroup shows lower initial scores and a slower growth rate. Note that these subgroup labels—HBFG and LBSG— indicate relative differences in intercepts and slopes between the two trajectories as identified through LCGA, without implying absolute measures of growth rate or baseline motor scale. In the HBFG subgroup (n=18), 10 of the infants were FT and 8 were PT. In the LBSG subgroup (n= 16), 9 of the infants were FT and 7 were PT. The LBSG subgroup has an average initial raw BSID gross motor at 11 (SD: 4) and a growth rate of 3 across the monthly visits, and HBFG has an average initial raw BSID gross motor at 12 and a growth rate of 5 across the monthly visits. The last-visit average gross motor scores were again higher in the HBFG subgroup at 24, with the LBSG at 19. While infant data used in this study showed a range of study entry/exit points, the distribution of data for each month (from 1-6) were similar between both subgroups. The adjusted age was 145 days, with a range from 37-276 in LBSG, followed by HBFG (140 days, Range: 40-276). Sex differences were 12/18 (67%) female in HBFG and 10/16 (62.5%) female in LBSG.

**Figure 4.**
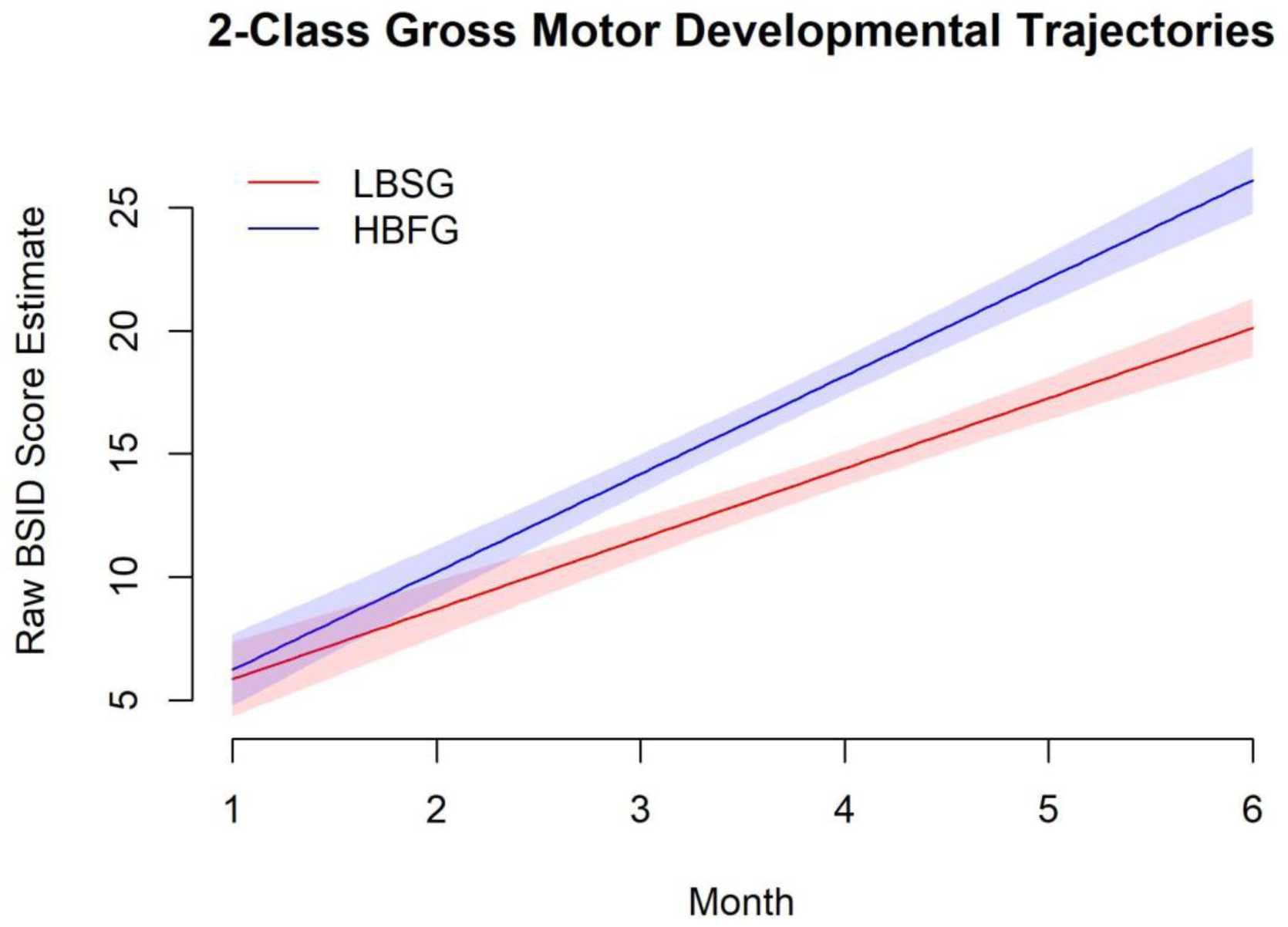
Gross motor developmental trajectories as defined by latent class growth analysis. Shaded areas represent 95% CI. LBSG = low-baseline slow-growth subgroup. HBFG = high-baseline fast-growth subgroup. X-axis is age in months, adjusted for infants born preterm.

The mean raw gross motor score was highest in HBFG (17, SD: 6), followed by LBSG (14.65, SD:4.39). The mean fine motor score was highest in LBSG (11, SD:6), followed by HBFG (11 SD: 5). It is important to note that although the raw motor scores may be lower in one group than the other, these scores are not adjusted to reflect age, although age distribution was similar between both subgroups. For this purpose, we also included the BSID Motor Composite Score, as described in the introduction.

The standardized score was highest in HBFG (101.03, SD: 13.54), followed by LBSG (89.45, SD: 12.60). Other demographic and visit information for each class can be found in **Table 3**.

**Table 3.**
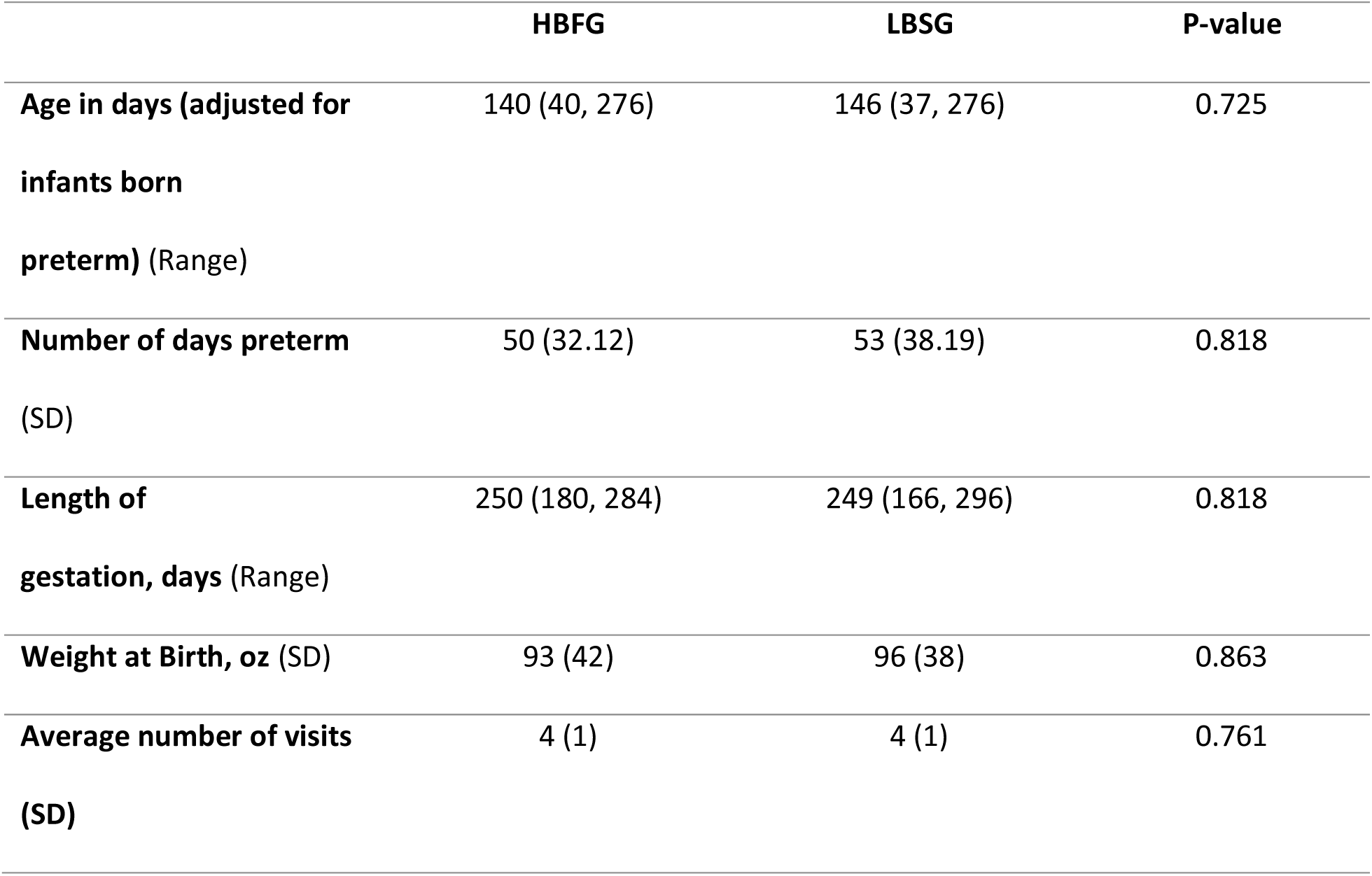
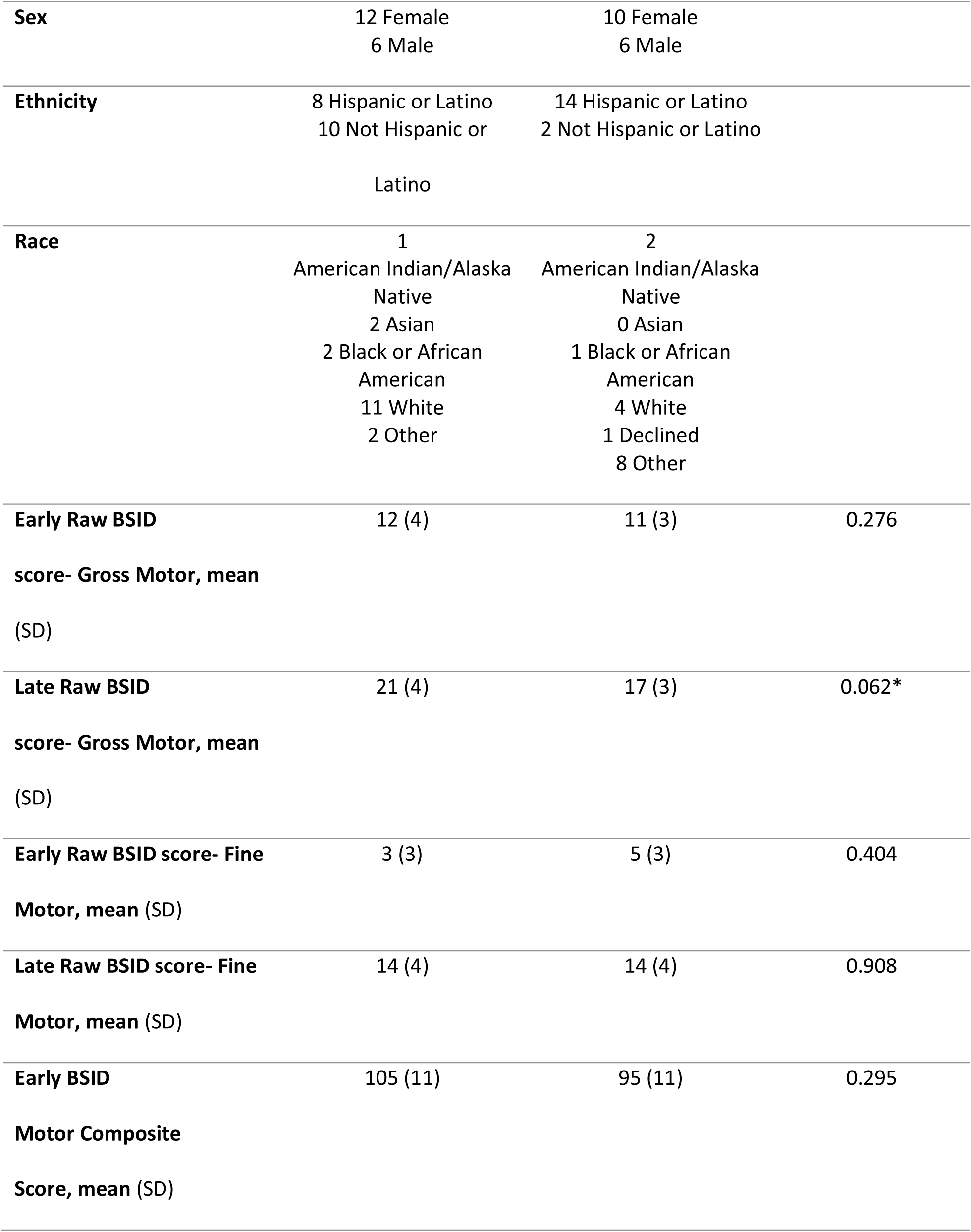
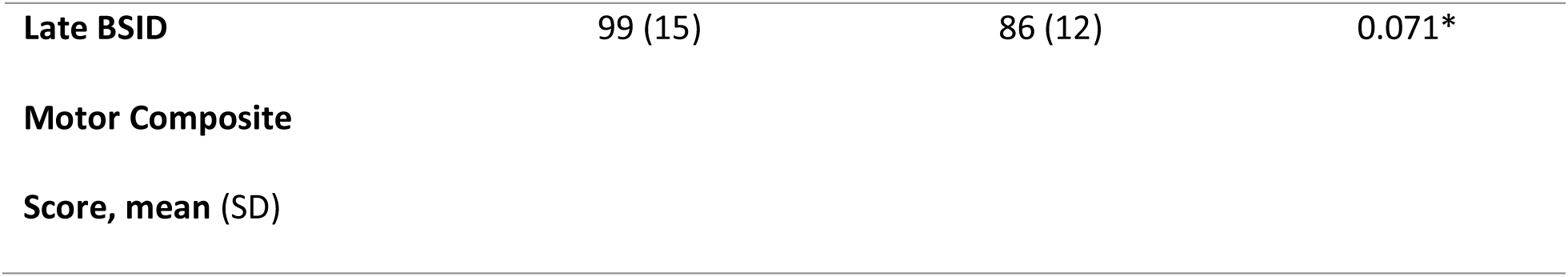
Demographic and motor assessment information for the gross motor linear, 3-class latent class growth analysis (LCGA) model. Demographic and motor assessment information for the gross motor linear, 3-class latent class growth analysis (LCGA) model. BSID = Bayley Scales of Infant Development III. Early scores are calculated based on data from infants aged 1-3 months; late scores are calculated based on data from infants aged 4-6 months. Means are listed with standard deviation (SD) or range (Range) in parenthesis. P-values calculated using Wilcoxon Rank Sum Test. LBSG = low-baseline slow-growth subgroup. HBFG = high-baseline fast-growth subgroup. * = p <0.10

**Table 4.**
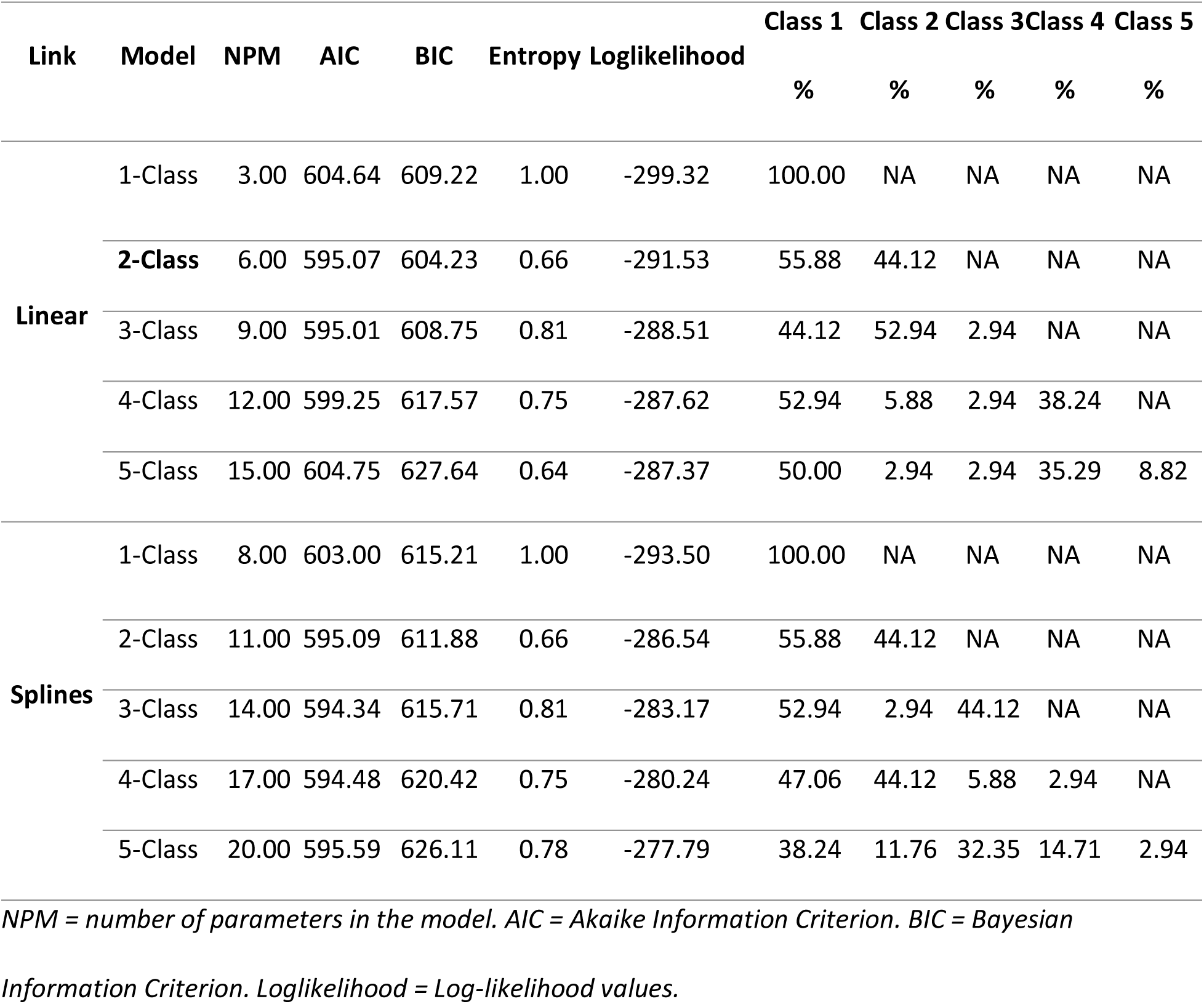
Latent Class Growth Analysis results for a) gross motor and b) fine motor based on class and link. Latent Class Growth Analysis results for a) gross motor and b) fine motor based on class and link. NPM = number of parameters in the model. AIC = Akaike Information Criterion. BIC = Bayesian Information Criterion. Loglikelihood = Log-likelihood values.

| Link | Model | NPM | AIC | BIC | Entropy | Loglikelihood | Class 1 | Class 2 | Class 3 | Class 4 | Class 5 |
| --- | --- | --- | --- | --- | --- | --- | --- | --- | --- | --- | --- |
|  |  |  |  |  |  |  | % | % | % | % | % |
| **Linear** | 1-Class | 3.00 | 604.64 | 609.22 | 1.00 | -299.32 | 100.00 | NA | NA | NA | NA |
|  | 2-Class | 6.00 | 595.07 | 604.23 | 0.66 | -291.53 | 55.88 | 44.12 | NA | NA | NA |
|  | 3-Class | 9.00 | 595.01 | 608.75 | 0.81 | -288.51 | 44.12 | 52.94 | 2.94 | NA | NA |
|  | 4-Class | 12.00 | 599.25 | 617.57 | 0.75 | -287.62 | 52.94 | 5.88 | 2.94 | 38.24 | NA |
|  | 5-Class | 15.00 | 604.75 | 627.64 | 0.64 | -287.37 | 50.00 | 2.94 | 2.94 | 35.29 | 8.82 |
| **Splines** | 1-Class | 8.00 | 603.00 | 615.21 | 1.00 | -293.50 | 100.00 | NA | NA | NA | NA |
|  | 2-Class | 11.00 | 595.09 | 611.88 | 0.66 | -286.54 | 55.88 | 44.12 | NA | NA | NA |
|  | 3-Class | 14.00 | 594.34 | 615.71 | 0.81 | -283.17 | 52.94 | 2.94 | 44.12 | NA | NA |
|  | 4-Class | 17.00 | 594.48 | 620.42 | 0.75 | -280.24 | 47.06 | 44.12 | 5.88 | 2.94 | NA |
|  | 5-Class | 20.00 | 595.59 | 626.11 | 0.78 | -277.79 | 38.24 | 11.76 | 32.35 | 14.71 | 2.94 |

When comparing the HBFG and LBSG subgroups in terms early (monthly age 1∼3) scores and late (monthly age 4∼6) scores, it shows a marginally significant difference (0.05 < p < 0.10) between the late gross motor raw scores (p = 0.062) and between the late motor composite scores (p = 0.071).

### Phenotyping Developmental Trajectories of Fine Motor Scales

For the fine motor models, the linear, 2-class model was chosen as the best fit. The AIC and BIC values reported were 595.07 and 604.23, respectively. The linear model also had lower AIC and BIC values than their nonlinear counterparts. The 3-class model presents comparable statistical measures as the 2-class model but was not selected due to the sole participant being delineated into the third class. In the linear 2-class fine motor skill model, 55.88% of participants were designated to Class 1 (HBFG), and 44.12% to Class 2 (LBSG).

Developmental trajectories of each class can be found in **Fig. 5**. In LBSG (n=15), 6 of the infants were FT and 9 were PT. In HBFG (n= 19), 13 of the infants were FT and 6 were PT. The HBFG subgroup has an average initial raw BSID fine motor at 7 (SD: 4) and a growth rate of 4 across the monthly visits, and the LBSG subgroup has an average initial raw BSID fine motor score at 5 (SD: 3) and a growth rate of 4 across the monthly visits. The last visit’s average fine motor also differed, with HBFG at 17 and LBSG at 16. The adjusted age was highest in LBSG (151 days Range: 65-276), followed by HBFG (129 days Range: 37-215. The sex differences were 9/15 (60%) female in LBSG 13/19 (68%) female in HBFG.

**Figure 5.**
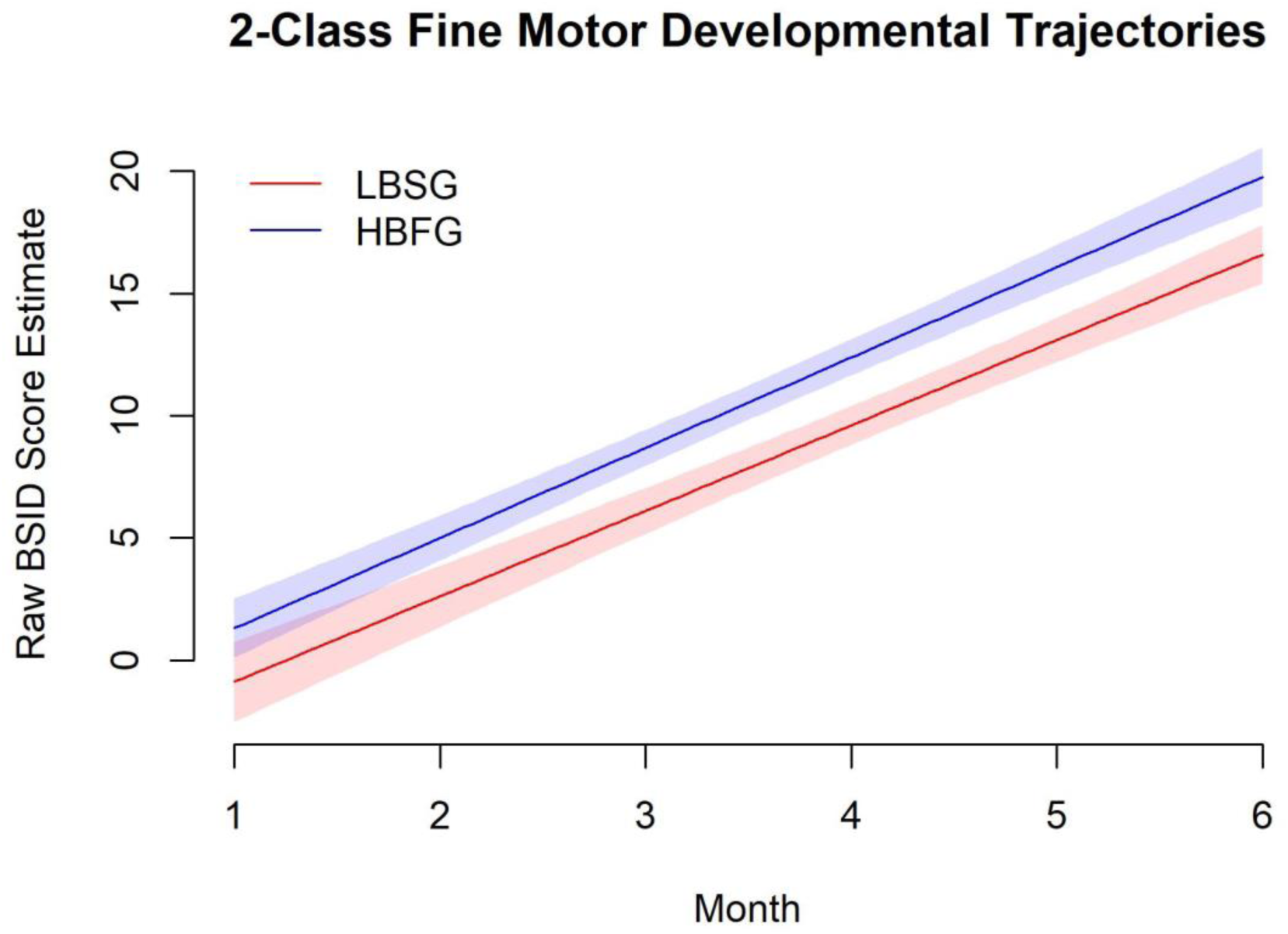
Fine motor developmental trajectories as defined by latent class growth analysis. Shaded areas represent 95% CI. LBSG = low-baseline slow-growth subgroup. HBFG = high-baseline fast-growth subgroup. X-axis is age in months, adjusted for infants born preterm.

The mean raw gross motor score was highest in HBFG (12, SD: 6), followed by LBSG (10, SD: 5). The mean fine motor score was highest in HBFG (16, SD: 6), then LBSG (16, SD: 5). Again, these scores are not adjusted to reflect age. The standardized Motor Composite score was highest in HBFG (100.72, SD: 13.94), then LBSG (89.02, SD: 11.96). Other demographic and visit information for each class can be found in **Table 5**. The Wilcoxon Rank Sum Test, as established in the Methods section, was conducted between both subgroups in terms early (monthly age 1∼3) scores and late (monthly age 4∼6) scores. A significant difference (p < 0.05) was found between the late motor composite scores between both groups (p = 0.04).

**Table 5.**
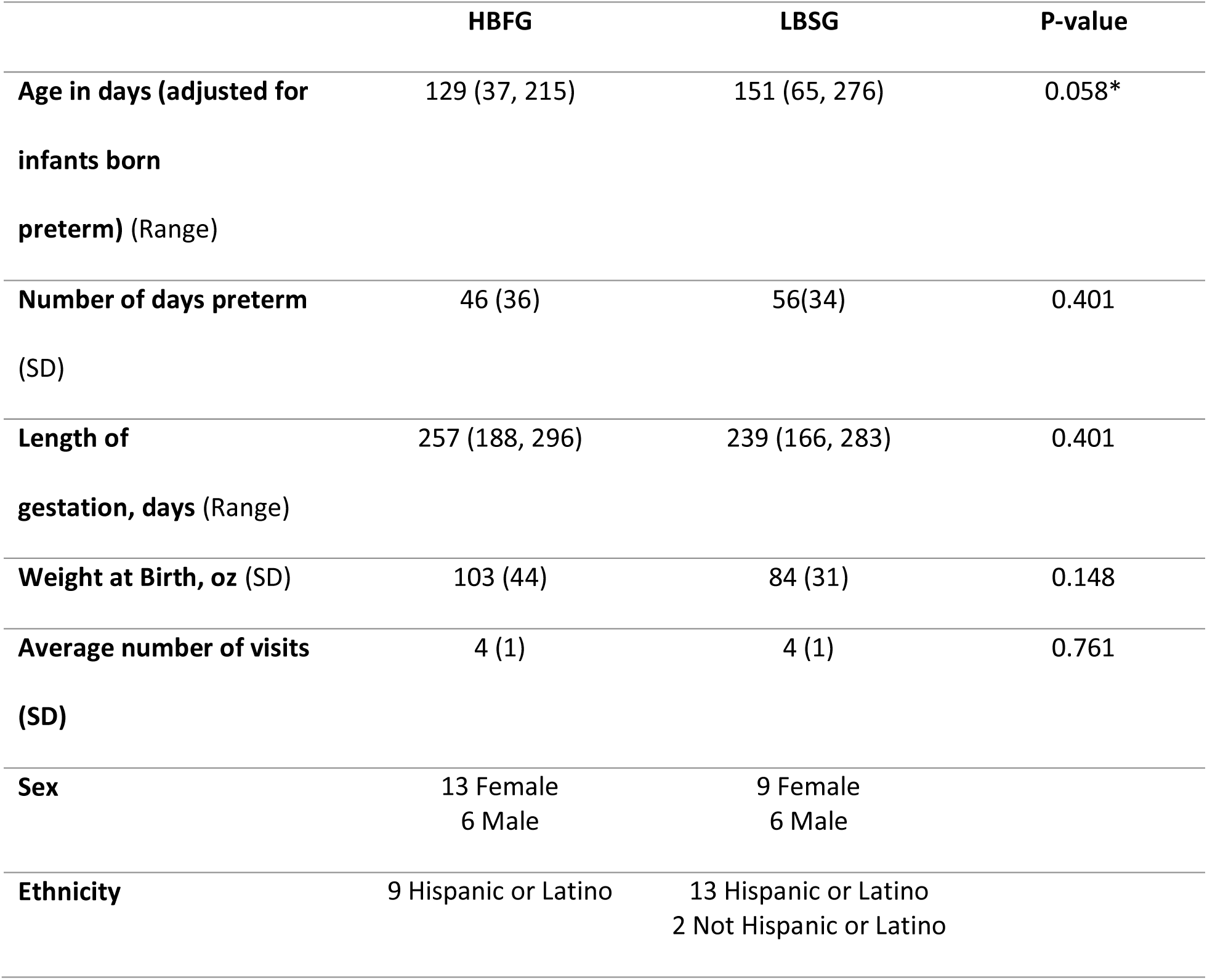
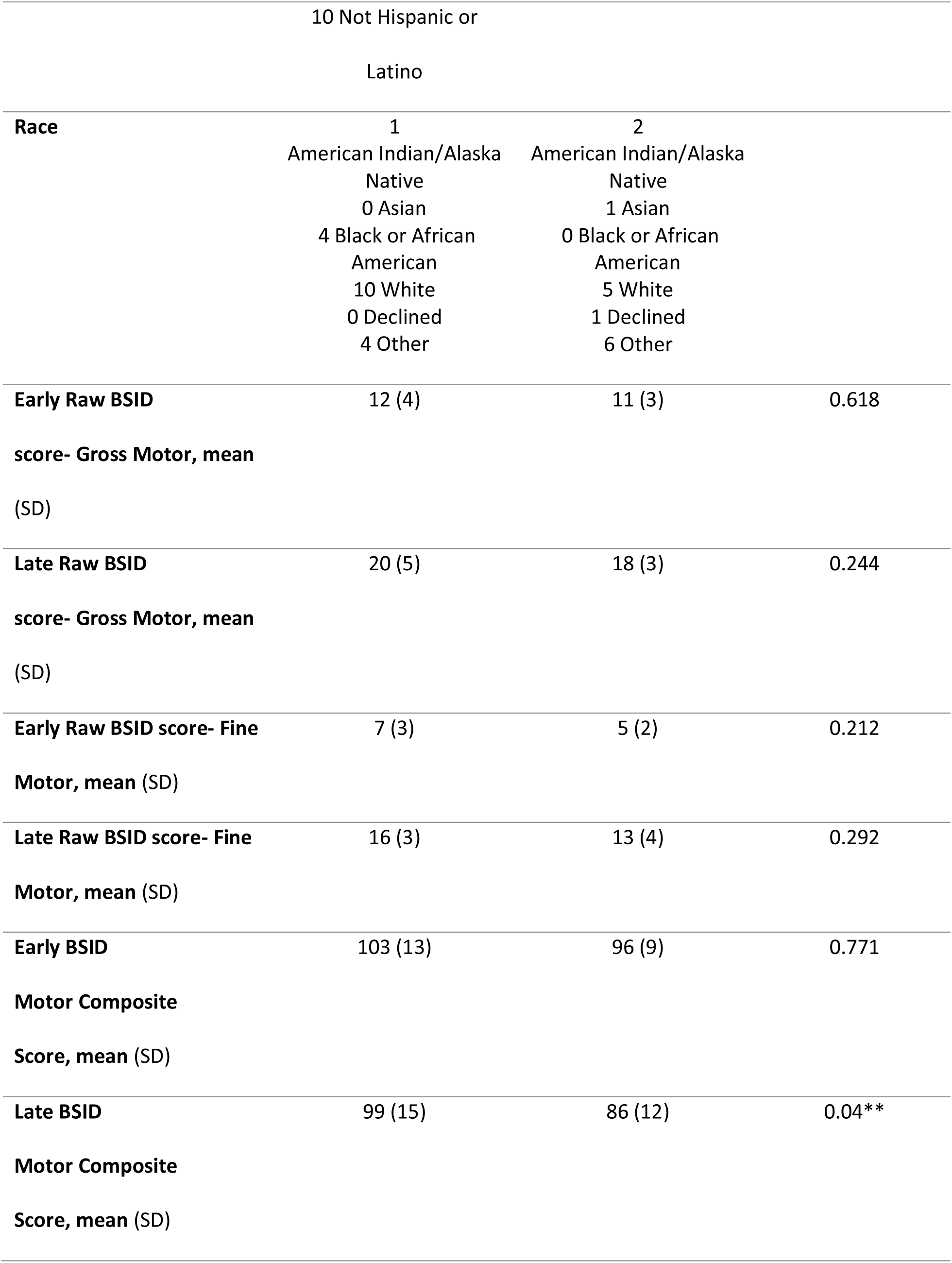
Demographic and motor assessment information for the fine motor linear, 3-class latent class growth analysis (LCGA) model. Demographic and motor assessment information for the fine motor linear, 3-class latent class growth analysis (LCGA) model. BSID = Bayley Scales of Infant Development III. Early scores are calculated based on data from infants aged 1-3 months; late scores are calculated based on data from infants aged 4-6 months. Means are listed with standard deviation (SD) or range (Range) in parenthesis. P-values calculated using Wilcoxon Rank Sum Test. LBSG = low-baseline slow-growth subgroup. HBFG = high-baseline fast-growth subgroup. * = p <0.10. ** = p <0.05. BSID = Bayley Scales of Infant Development III. Early scores are calculated based on data from infants aged 1-3 months; late scores are calculated based on data from infants aged 4-6 months. Means are listed with standard deviation (SD) or range (Range) in parenthesis. P-values calculated using Wilcoxon Rank Sum Test. LBSG = low-baseline slow-growth subgroup. HBFG = high-baseline fast-growth subgroup. * = p <0.10. ** = p <0.05.

LCGA estimates the probability of belonging to each class based on observed data patterns rather than deterministically assigning individuals. When evaluating infants’ class membership probabilities for fine and gross motor skills, 38% of infants (13 out of 34) showed varying classifications, with the highest likelihood shifting between the LBSG and HBFG groups, or vice versa.

## Discussion

This study explores the benefit of using unsupervised machine learning, i.e., LCGA analysis, to characterize motor development trajectories across infants during the first half-year of life. It emphasizes the heterogeneity within populations while elucidating that distinct developmental tracts may provide insight into targeted clinical care. Ultimately, the linear, 2-class model was selected for both the gross and fine motor models. Comprehensive consideration is necessary for choosing the suitable class number for the analysis. This includes consideration of multiple statistical measures, which provide a more holistic evaluation of the phenotyping results after LCGA. Additionally, model complexity and sample distribution need to be factored in given the limited sample size available in this type of research to mitigate overfitting and foster model generalization to unseen data. In this study, although a higher-class number sometimes offers a slight improvement in statistical measures, those are offset by the model complexity and sample spread as compared to the 2-class model. This view also emphasizes the necessity of larger data pools to extract additional patterns that may emerge, as further discussed in the limitations section below.

Additionally, the groupings between the two classes were not the same when comparing the gross motor and fine motor skill developmental trajectories. This suggests that, while evidence of the correlation between gross motor and fine motor skills exists (Gonzalez et al., 2019), and they are not independent from one another, nor are they interchangeable. At a groupwise level, developmental trajectories from both fine and gross motor may yield different patterns across infants. Individually, this indicates that some infants may have different developmental trajectories for fine and motor scales.

This is corroborated by the fact that 38% (13 out of 34) of infants in our study switched class membership (from LBSG to HBFG, or vice versa) in fine and gross motor developmental trajectories. How these separate trajectories may correlate with later cognitive development shows is still being elucidated (Piek et al., 2008). While this study does not aim to predict the outcomes of infants, it provides evidence that subdividing developmental skillsets may be ideal for creating individualized support. Additionally, the Wilcoxon Rank Sum Tests within both motor categories provide evidence that a single risk factor may not exclusively determine the categorization of the infant’s motor trajectory, including pretermness. Across most characteristics, the subgroups revealed significant heterogeneity.

Notably, the fine motor subgroups demonstrated a marginal significance between ages of the infants, with the LBSG older than the HBFG. These claims, however, are restricted due to the sample size of the study, which will be discussed further in the limitations section. Future work is necessary to investigate the heterogeneity of developmental tracts of infants and incorporate this into a predictive modeling system.

As expected, the raw scores of all groups increased as infants age, indicating that infants were progressing in their ability to complete fine and gross motor tasks as they age. Both the gross motor and fine motor models appear to have distinctions within developmental trajectories. One of the classes has a higher trajectory than the other. The High Baseline Fast Growth subgroup was named based on the relatively higher rates of growth and scoring over time. The Low Baseline Slow Growth subgroup had relatively lower rates of growth and lower score values based on the motor subscale of interest (gross or fine). The exception to this is that the fine motor had a slightly higher rate of growth in the LBSG subgroup than the HBFG. This could be the result of a small sample size and the significantly lower starting scores for the group. Additionally, the later (grouped as months 4, 5, and 6) BSID motor composite scores were significantly different in both the fine motor and gross motor models as supported by the Wilcoxon Rank Sum Test. The gross motor model also demonstrated a difference between the later raw gross motor scores, though this effect was not reciprocated in the fine motor model. These differences provide evidence for distinct trajectories able to be extracted from the growth models.

Overall, one class in both the fine and gross motor models included a majority of infants born FT, one class a majority of infants born PT, and one class with an infant born FT that scored much higher than others. It is interesting to note that although there was a majority of one group within a class, there existed notable variability and heterogeneity across classes. This is consistent with literature that identifies preterm as a risk factor for developmental delay or disorder (Bos et al., 2013; Hee Chung et al., 2020; Kerstjens, Bocca-Tjeertes, et al., 2012). However, this work also provides further evidence for heterogeneity within populations, both full-term and pre-term, which is also supported by neurological and developmental science (Burstein et al., 2024; Dimitrova et al., 2020). By subdividing a population by developmental trajectory as opposed to risk factor, we have a useful baseline for identifying differences and characteristics of group patterns. This also supports the reasoning to include full-term infants in the analysis. While infant development assessments, such as the BSID, are normally reserved for individuals who are at risk of developmental disability, understanding typical developmental trajectory and its patterns better equips clinicians and caregivers to provide individual intervention and support when needed. Perhaps the key to identifying those with the highest need of intervention will benefit from “complexity” or utilizing a multifactorial approach and avoiding a reduction to one or two risk factors (D’Souza et al., 2017).

Developmental trajectory is a complex and multifaceted field of study. Many factors could contribute to the developmental trajectory, including 1) biological or environmental factors, and 2) the impact of treatment or intervention on the trajectory and the resultant subgroup membership, which warrant future research to untangle and may be better elucidated through alternative machine learning approaches. This study does not serve to inflexibly categorize infants into a predestined trajectory; rather, it serves to retroactively analyze whether patterns emerge within infant subpopulations, independent of gestational age at birth or other predetermined details. This study was effectively delineated by data-driven analyses, wherein only the BSID scores served as a differentiating factor. We see evidence of this in the heterogeneity of each class. While this limits the ability to predict or draw conclusions about potential risk factors, it does include potential avenues for future research questions.

### Limitations

Despite patterns and insights found in the study, several limitations must be addressed. Due to the secondary analysis nature of the study, a significant limitation is the infant sample size (n=34 after exclusions). This challenge is common in developmental studies, particularly when collecting longitudinal samples from infant participants, which requires many resources and often contains missing data. This leads to a field-wide concern of data replication and robustness. These limitations are elucidated by Davis-Kean & Ellis (Davis-Kean & Ellis, 2019). These authors emphasize the importance of delineating between confirming and exploratory research. Additionally, meta-analyses highlight both the prevalence and the cautions associated with reporting marginal significance (i.e., 0.05 < p < 0.10). Following these guidelines, we reaffirm that the primary goal of this study is to uncover and illustrate the diverse developmental trajectories in infants, considering factors beyond preterm birth. The Wilcoxon Rank Sum Tests serve as an exploration of differences in characteristics between subgroups to inform future research questions. The findings—particularly the marginal significance observed in early and late gross motor scores between latent subgroups—require further validation through future studies with larger sample sizes. Ultimately, the Wilcoxon Rank Sum Tests reinforces the heterogeneity between subgroups and warns against the oversimplification in delineating motor development trajectory.

Because of the limited number of participants, there is also a potential risk of overfitting the model to the infants. This was especially the case in the 3-class models and above. Although the 2-class model was chosen based on statistical criteria, as discussed, a larger dataset would increase the statistical power and further validate the findings. Additionally, infants with unstable medical conditions were excluded from the original studies. This also may skew the potential developmental trajectories.

For this reason, our results cannot directly inform clinical care or provide individualized predictive properties. Additional studies with larger sample sizes with a widened range of developmental status at the time of study inclusion are warranted to increase the statistical power of the models. However, conducting an unsupervised learning approach, such as LCGA, provides insights and foundational information for future research questions.

Another limitation is the use of the BSID to assess monthly developmental trajectory. The purpose of the BSID was not created to measure change over time; it is meant to provide a relative indication of the infant development at a point in time. In a clinical setting, the BSID is generally not measured monthly. According to the BSID-III Manufacturer, “An interval of approximately 3 months is recommended for children under 12 months of age; an interval of approximately 6 months is recommended for children older than 12 months, although shorter intervals can be used if warranted.” These intervals are generally created to prevent practice or learning effects. However, the BSID tasks for young infants are mostly based on observation of typical behaviors as opposed to tasks where repeated assessment would be influenced by learning. For example, eyes following a moving ring is a task for young infants, as is head control while tilting the body at different angles and types of sounds infants are able to make. Additionally, caution about drawing longitudinal interpretations of BSID scales from adjacent time points at an individual level is warranted. The composite scores are dependent on normalized references of a population in time. For this reason, we identified the raw gross and fine motor scores instead of the composite score as the primary determinant of the model. Utilizing the raw and fine motor scores allows for 1) separating the predictive models based on either fine or motor development, and 2) provides a stable measure of growth over time, irrespective of age.

Conducting a secondary analysis yields limitations to the type of patterns that can be identified by this model. While a number of demographics were available to compare between subgroups, there are many more that could provide further insight into individual growth trajectories. For example, socio-economic status, quality of caregiver-infant interaction, and genomic data were not analyzed in this study and may create more robust developmental models. Additionally, while the LCGA is ideal for elucidating trajectories within a heterogeneous population, it is incapable of determining individual trajectory prediction or risk factor influence. Future studies may use this analysis as a precursor to more comprehensive research questions and models.

## Data Availability

All data produced in the present study are available upon reasonable request to the authorsf

